# Efficacy and safety of 10-day versus 14-day bismuth-containing quadruple therapy for H. pylori eradication: A systematic review and meta-analysis

**DOI:** 10.1101/2024.08.18.24312061

**Authors:** Najam Gohar, Zoya Ejaz, Faizan Ahmed, Abdul Rafay, Abdullah Humayun, Momna Nisar, Ali Mushtaq, Aanusha Ghouri, Fatima Zafar, Hira Khalid, Sania Afzal, Hammad Khan, Huzaifa Ahmed Cheema, Muhammad Shahzil, Essam Rashad, Rehmat Ullah Awan, Prasun K Jalal

## Abstract

**Background:** Nearly half of the world population is infected by *Helicobacter pylori (H. pylori).* Bismuth-containing quadruple therapy (BQT) has shown favorable outcomes. This study compares 10-day and 14-day BQT regimens to evaluate their efficacy, safety, and compliance rates.

**Methods:** We searched electronic databases from their inception until May 2024 to retrieve all randomized controlled trials (RCTs) that compared 10-day and 14-day BQT regimens for *H. pylori* eradication. Meta-analysis was performed using Review Manager 5.4. Dichotomous outcomes were compared using risk ratio (RR).

**Results:** Seven RCTs and a total of 2,424 patients were included in the meta-analysis. There was no significant difference in the intention-to-treat eradication rate (RR 0.97; 95% CI 0.94, 1.01) and the per-protocol eradication rate (RR 0.96; 95% CI 0.93, 1.00) between the 10-day BQT and 14-day BQT groups. Commonly reported adverse events in both groups were epigastric pain and discomfort, nausea, and vomiting. There was no significant difference in the risk of adverse events between the two groups (RR 0.80; 95% CI 0.63, 1.02). There was no significant difference in the compliance rate between the two groups (RR 1.02; 95% CI 1.00, 1.04).

**Conclusion:** The eradication rates, risk of adverse events, and compliance rates were comparable between the two groups. Future research comparing similar drug doses with larger sample sizes and longer patient follow-ups can improve the quality of results.

**Highlights:**

- Our meta-analysis comprising 2424 patients showed that patients receiving 10-day bismuth-containing quadruple therapy had comparable eradication rates to those receiving 14-day bismuth-containing quadruple therapy for *Helicobacter pylori*.
- Additionally, there was no significant difference between the two groups for compliance and risk of adverse events.
- Antibiotic resistance was associated with lower eradication rates in both treatment groups.

## Introduction

A significant proportion of the world’s population is afflicted by *Helicobacter pylori* (*H. pylori*) infection. In 2015, around 4.4 billion people worldwide were reportedly impacted by this infection [1]. The burden of H. pylori infection is higher in developing countries than in developed countries, with the highest prevalence in African and Eastern Mediterranean regions. The adult population Is affected by *H. pylori* infection more than children and adolescents, with a prevalence rate of 43.9% and 35.1% respectively [2]. *H. pylori* is recognized as the causative agent of various gastrointestinal conditions, including chronic gastritis, peptic ulcers, and gastric mucosa-associated lymphoid tissue (MALT) lymphoma [3,4].

Untreated *H. pylori* infection can result in accelerated loss of specialized glands in the stomach, leading to atrophic gastritis and increasing the risk of gastric carcinoma. Seeking timely effective treatment is crucial in preventing detrimental outcomes [5]. Elimination of infection is cost-effective, especially for high-risk individuals. The treatment costs in patients with successful eradication were found to be almost half that accrued by the individuals with treatment failure and associated complications [6].

Triple or quadruple therapy are the two commonly employed regimens for eradicating infection. Triple therapy includes a combination of a proton pump inhibitor (PPI) and two antibiotics, such as amoxicillin and clarithromycin or metronidazole. The efficacy of triple therapy is declining due to antibiotic resistance [7]. Quadruple therapy, which consists of adding bismuth to the triple therapy, is effective in areas with high antibiotic resistance [8]. In a meta-analysis, it was found that quadruple therapy has better cure rates than triple therapy [9].

The duration of bismuth quadruple therapy is still under evaluation. The previous systematic review and meta-analysis comparing the two treatment regimens exhibited comparable eradication rates, with a lower risk of adverse events in the 10-day bismuth-containing quadruple therapy (BQT) group. However, since the previous meta-analysis, three RCTs [10–12] have been published that have conflicting outcomes to one another and have not been incorporated into a meta-analysis, with one being the largest trial comparing the two regimens to date [10]. The new studies collectively comprise a sample size greater than the previous meta-analysis. A shorter duration of therapy can improve patient compliance, with potential savings in healthcare expenses and fewer drug shortage concerns.

This meta-analysis aimed to assess the effectiveness of 10-day BQT compared to 14-day BQT and to profile the adverse events associated with both treatment durations.

## Materials and Methods

This systematic review was conducted according to the methodological guidelines outlined in the Cochrane Handbook for Systematic Reviews of Interventions. The reporting follows the Preferred Reporting Items for Systematic Reviews and Meta-Analysis (PRISMA) statement [13]. The review protocol was registered with the International Prospective Register of Systematic Reviews (PROSPERO) under the identifier CRD42024551033. Ethical approval was not required for this study.

### Eligibility Criteria

The inclusion criteria were as follows: 1) study design: RCTs; 2) population: adults over 18 years of age, undergoing medical therapy for *H. pylori* eradication; 3) intervention: 10-day BQT therapy (quadruple therapy was defined as a combination of three medications including a PPI, and two antibiotics with any formulation of bismuth); 4) comparator: 14-day BQT therapy; 5) outcomes: reporting any outcome of interest.

The exclusion criteria were: 1) study designs other than RCTs, such as case series and case reports, quasi-randomized trials, and observational studies; 2) treatment regimens other than 10 and 14 days BQT; 3) treatment regimen not having bismuth in it; 4) studies conducted on animals; and 5) studies in languages other than English.

### Information Sources

A comprehensive search strategy was employed to identify relevant studies. We searched electronic databases and international trial registries from inception to May 2024. The search was conducted using the Cochrane Central Register of Controlled Trials (via The Cochrane Library), MEDLINE (via PubMed), Embase (via Ovid), ClinicalTrials.gov, and the World Health Organization International Clinical Trials Registry Platform (ICTRP) portal. No language restrictions were applied. Additionally, grey literature sources such as ProQuest Dissertations & Theses Global and OpenGrey were explored for potentially relevant data. Reference lists of included articles and relevant systematic reviews were hand-searched to identify further eligible studies. Forward citation tracking was conducted using the Web of Science to retrieve any additional pertinent research. A detailed search strategy utilizing keywords and Medical Subject Headings (MeSH) terms related to 10 and 14-day bismuth-containing quadruple therapy, *Helicobacter pylori*, and eradication is provided in **Supplementary Table 1.**

### Selection Process

A two-stage screening process was employed for article selection. All retrieved citations were uploaded to Rayyan AI, a web-based platform for systematic reviews. Two authors independently screened titles and abstracts against pre-defined eligibility criteria. Next, full-text articles of potentially eligible studies were retrieved and independently reviewed by the same authors. Disagreements were resolved through arbitration by a senior author.

### Data Collection Process and Data Items

After the process of study selection, data were extracted by two reviewers into a pre-piloted Microsoft Excel spreadsheet to ensure consistency of data extraction. Relevant data items were extracted, including patient characteristics (number of participants, their mean age in years, percentage of male population, confirmatory diagnosis for *H. pylori* infection, treatment-naive population), intervention details (including study arms and regimen), comparator details (14-day BQT regimen), study characteristics (trial name, first author, year of publication, name of country in which the study was conducted, duration of follow-up), and the outcome variables. Our primary outcomes were the intention-to-treat eradication rate and the per-protocol eradication rate. The intention-to-treat analysis (ITT) includes all participants who were randomized at the beginning of the study, irrespective of whether they completed the prescribed treatment or follow-up. Per-protocol (PP) only considers those participants in the analysis who completed the therapy as described in the protocol, without any major violations. Secondary outcomes included the incidence of adverse events and the compliance rate.

### Risk of Bias Assessment

Two authors assessed the risk of bias in the included studies using the revised Cochrane Risk of Bias tool for randomized trials (RoB 2.0) [14], which assesses bias in the following 5 domains: 1) bias arising from the randomization process; 2) bias caused by deviations from intended interventions; 3) bias caused by missing outcome data; 4) bias in the measurement of the outcome, and 5) bias in the selection of the reported result. Two authors independently rated the risk of bias for each included study as low, high, or some concerns. A third author resolved any disagreement between them.

### Data Synthesis

We used Review Manager (RevMan, version 5.4; The Cochrane Collaboration, Copenhagen, Denmark) for statistical analysis. Dichotomous outcomes were reported as relative risk (RR) with 95% confidence intervals (CIs). The random-effects model with the Mantel-Haenszel method was used to perform meta-analyses. A two-tailed p-value less than 0.05 was taken to be significant.

For each synthesis, the I^2^ index and the chi-square test were used for the assessment of heterogeneity, and a *p*-value of <0.1 was considered significant for the heterogeneity of the included studies.

## Results

The study selection process is illustrated in the PRISMA flowchart in **Figure 1**. An initial search yielded 309 studies. After removing duplicates, screening titles, and abstracts, the studies were assessed by their full-text manuscripts. Subsequently, seven RCTs were included in the analyses. **Table 1** summarizes the main characteristics of all included studies, such as age, country, and treatment drugs. Seven RCTs [10–12,15–18] of BQT were analyzed to compare the eradication rates of treatment durations of 10 and 14 days. All studies reported both ITT and PP eradication rates for the specified treatment durations. Additionally, one of the studies [16] had two pairs of intervention and control groups (10-day and 14-day bismuth-quadruple therapies and moxifloxacin-bismuth combined therapies), which were pooled separately in the meta-analysis. The studies also reported overall incidence of adverse effects and patient compliance. In the trials, 2424 patients were assigned to either the 10-day duration group (n = 1223) or the 14-day duration group (n = 1201). The trials were conducted in China (4 trials), Italy, Turkey, and Taiwan. All trials enrolled treatment naïve patients. Confirmation of *H. pylori* diagnosis was made by 13-C Urea breath test (13C-UBT), gastric biopsy and histology, and tissue culture. Detailed study characteristics are provided in the **supplementary table S2**.

**Figure 1:**
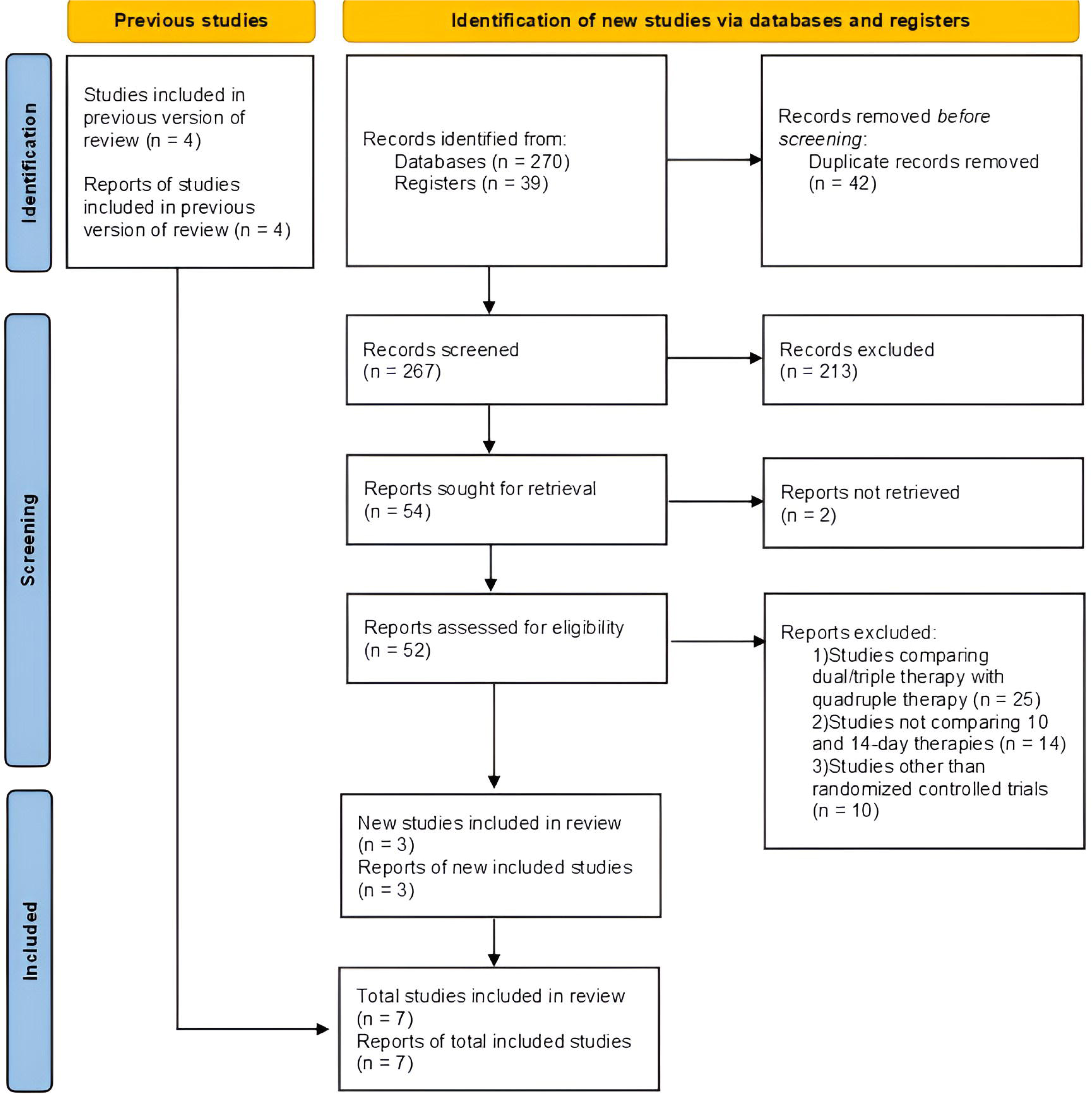
PRISMA flowchart for selection of studies.

**Table 1:** Study characteristics of the included studies.

| Study ID | Location | Included study arms | Included Participants | Mean Age ± SD in years | Male % | Duration of Follow-up |
| --- | --- | --- | --- | --- | --- | --- |
| **Chen 2020** | China | 10-day vs 14-day Rabeprazole BQT | 120 vs 120 | 45.89 ± 12.92 vs 48.11 ± 12.39 | 56.7 vs 50.8 | 4 weeks |
| **Dore 2011** | Italy | 10-day vs 14-day Bismuth, Tetracycline, Metronidazole, and Esomeprazole | 215 vs 202 | 52 vs 53 | 37.7 vs 35.6 | 4-6 weeks |
| **Etik Ozer 2019** | Turkey | 10-day vs 14-day bismuth-containing quadruple treatment group (10-BQT vs 14-BQT groups) | 54 vs 54 | 40 ± 11.0 vs 44 ± 12.5 | 31.5 vs 35.2 | 6-12 weeks |
| | | 10-day vs 14-day moxifloxacin-bismuth combined treatment groups (10-MBCT vs 14-MBCT groups) | 54 vs 54 | 41 ± 11.5 vs 43 ± 11.5 | 40.7 vs 42.6 | |
| **He 2023** | China | 10-day vs 14-day Antofloxacin containing BQT (ANT10 vs ANT14) | 395 vs 387 | 43.0 ± 11.8 vs 44.2 ± 11.1 | 51.6 vs 55.3 | 6 weeks |
| **Lu 2023** | China | 10-day vs 14-day Vonoprazan BQT | 78 vs 78 | 37.1 ± 19.7 vs 35.9 ± 11.5 | 41.0 vs 47.4 | 6-8 weeks |
| **Niu and Bai 2022** | China | 10-day vs 14-day Ilaprazole, Amoxicillin, Furazolidone, Bismuth Glycyrrhizinate (10-IAFB vs 14-IAFB) | 150 vs 150 | 44.25 ± 12.67 vs 44.10 ± 13.90 | 45.5 vs 42.1 | 4-6 weeks |
| **Yang 2024** | Taiwan | 10-day vs 14-day Bismuth Quadruple Therapy (10-BQT vs 14-BQT) | 157 vs 156 | 55.0 ± 14.6 vs 55.0 ± 13.0 | 52.9 vs 42.9 | At least 4 weeks |

### Risk of Bias Assessment

One study exhibited a low risk of bias, while there were some concerns of bias in the other six studies. These concerns mainly arose due to issues in the randomization process (two studies) and the selection of reported results (four studies). Additionally, one study had a considerable loss of population to follow up. The details of the bias assessment are provided in the **supplementary table S3**.

### Results of the meta-analysis

#### Eradication Rate

Seven studies reported data on the eradication rate. For the ITT analysis, the eradication rate for the 10-day group was 86.6%, while that for the 14-day group was 90.3%. There was no significant difference between the two treatment groups (RR 0.97; 95% CI 0.94, 1.01; I^2^=38%) (**Figure 2)**. In the PP analysis, the eradication rates were 90.8% and 96.0% in the 10-day and the 14-day groups, respectively, there was no statistically significant difference in the eradication rates of the 10-day and the 14-day groups (RR 0.96; 95% CI 0.93, 1.00; I^2^=75) in the PP analysis (**Figure 3)**.

**Figure 2:**
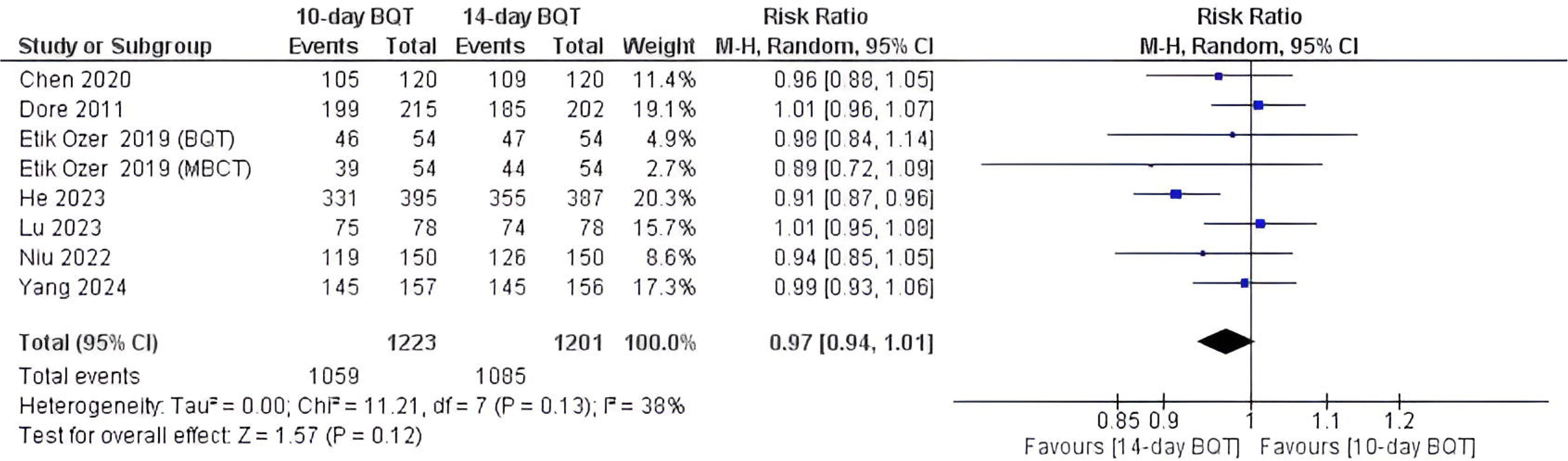
Forest plot for eradication (Intention-to-treat analysis).

**Figure 3:**
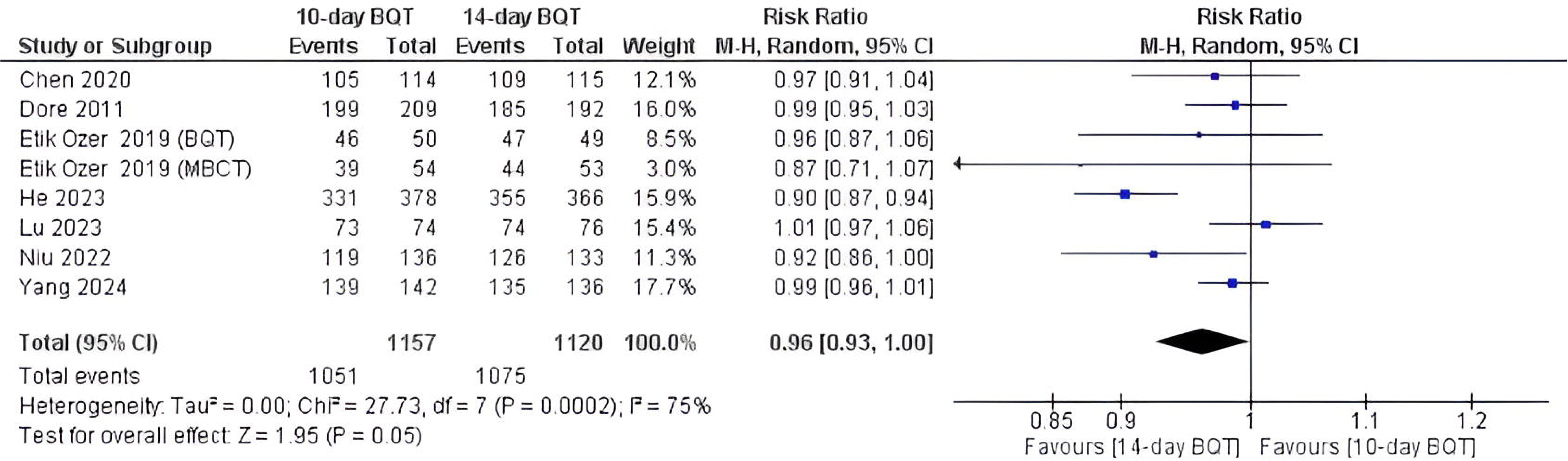
Forest plot for eradication rate (Per-protocol analysis).

#### Adverse Effects

All the included studies mentioned details regarding the adverse effects, with gastrointestinal symptoms being the most prominent ones. However, treatment duration did not significantly impact the risk of adverse events, as the risk of adverse events was not significantly different between the two groups (RR 0.80; 95% CI 0.63, 1.02; I^2^=59%) (**Figure 4**). The studies demonstrated significant heterogeneity.

**Figure 4:**
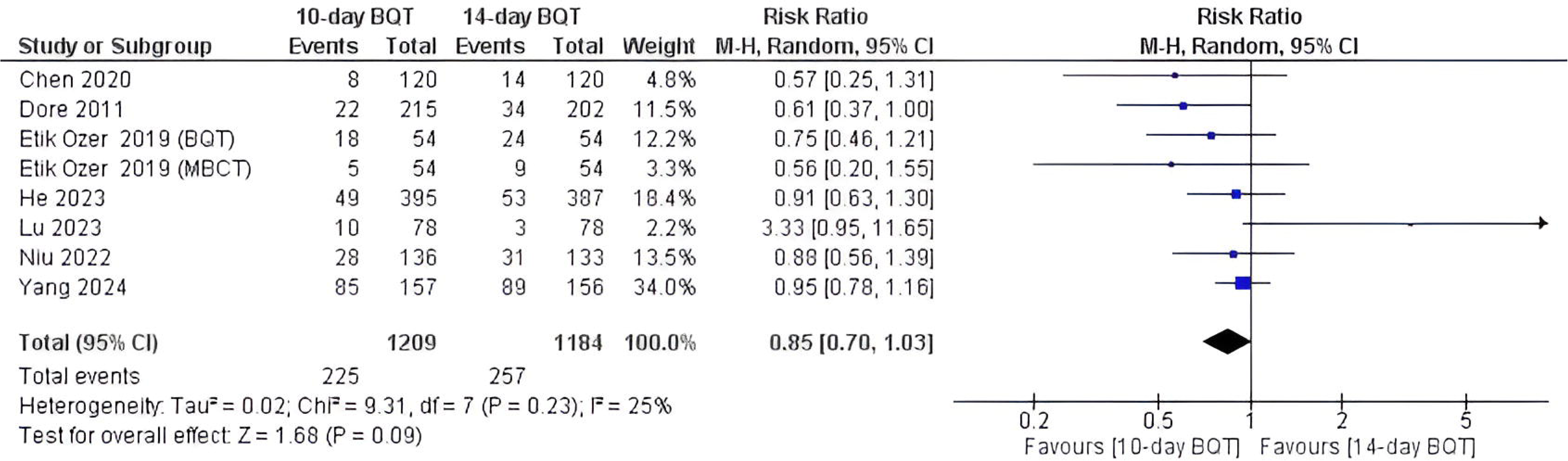
Forest plot for adverse events.

The adverse effects most commonly reported by studies were epigastric discomfort and pain, nausea, and vomiting. Additionally, the study by Yang et al. reported a significantly lower incidence of dizziness and vomiting in the 10-day group compared to the 14-day group [12].

#### Compliance

The compliance rate was reported by five studies included in the analysis. The compliance rate in the 10-day group was 94.6%, compared to 93.0% in the 14-day group. There was no significant difference in the compliance rate between the study groups (RR 1.02; 95% CI 1.00, 1.04; I^2^=0%) (**Figure 5**). There was no significant heterogeneity observed in the studies assessing for compliance rate.

**Figure 5:**
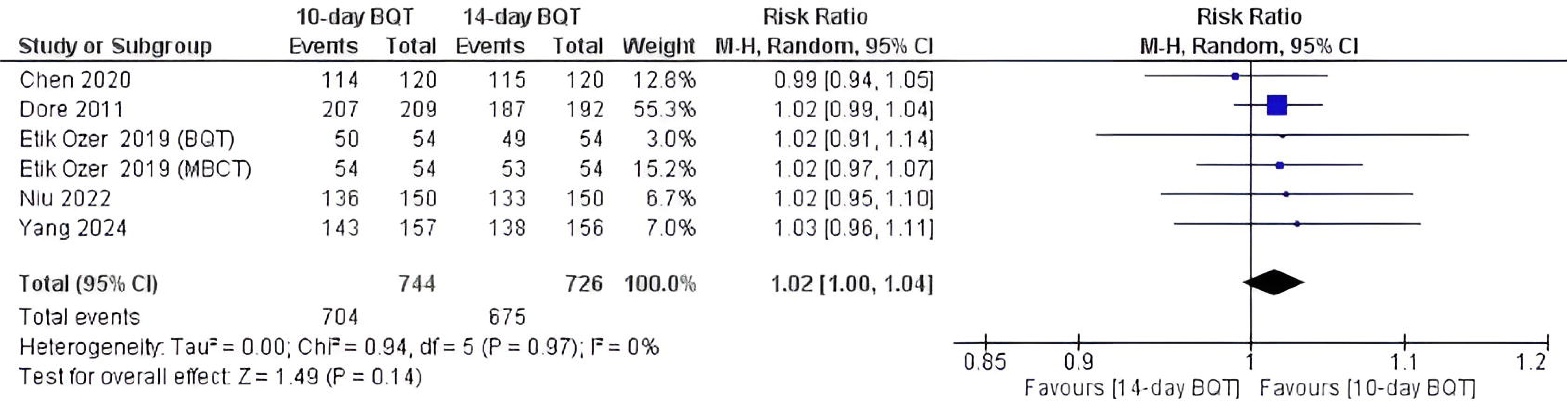
Forest plot for compliance rate.

## Discussion

In this meta-analysis, which included data from 7 RCTs, we evaluated the efficacy and safety of two treatment regimens for treatment-naive *H. Pylori*-infected individuals: a 10-day BQT and a 14-day BQT. The analysis included 2,424 patients and concluded that there was no significant difference in eradication rates between the two groups. Furthermore, the risk of adverse events and compliance rates were comparable between the two treatment durations. These findings suggest that both regimens are equally effective and safe for treating *H. Pylori* infections, providing flexibility in treatment duration without compromising the efficacy or safety. The risk of bias in the studies included in this review was mostly moderate, with one study having a low risk of bias.

Our findings align with previous studies showing no significant difference in eradication rates between 10-day and 14-day BQT regimens for *H. Pylori.* However, the previous meta-analysis assessing the two treatment regimens exhibited a statistically significant difference in the risk of adverse events, with the 10-day group having fewer adverse reactions [19]. This was not concordant with our meta-analysis, as the two groups demonstrated no significant difference in the risk of adverse events. This is further backed by the latest trials by He et al., Lu et al., and Yang et al. that showed comparable risk of adverse events between the two groups [10–12]. This difference can be explained by the observation that meta-analyses involving studies with small sample sizes may provide significant results that are not substantiated by subsequent larger trials [20].

Among the included studies, the trial by He et al. showed significant improvement in eradication rates in the 14-day BQT group over the 10-day BQT group in both the ITT and PP analysis. This study was also the largest trial to date comparing the two therapies. Drug resistances were demonstrated against clarithromycin and antofloxacin, with minimal resistance against amoxicillin. This study also demonstrated that drug resistance significantly reduced the odds of *H. pylori* eradication. The 10-day therapy group had a significantly lower eradication rate than the 14-day therapy group for antibiotic-resistant strains. [10]. Niu et al. demonstrated that the greatest incidence of resistance occurred against clarithromycin, metronidazole, and levofloxacin. Additionally, they demonstrated fewer costs incurred by patients on 10-day therapy compared to 14-day therapy [18]. Recent trials have also shown greater cost-effectiveness of the shorter-duration therapy compared to the longer-duration regimen [11].

Current guidelines advocate for the triple as well as bismuth-containing quadruple regimens. While the World Gastroenterology Organization (WGO) recommends a 7-day BQT, 10-day and 14-day regimens are also widely recognized and used, especially in areas of higher antibiotic resistance [21]. While metronidazole and tetracycline are the most preferred antibiotics, the addition of bismuth to standard triple therapy containing clarithromycin has shown favorable results [22]. There is current evidence to support that the 7-day therapy may not be inferior to 14-day therapy against antibiotic-resistant strains [23]. Further investigation into the treatment of drug-resistant strains is still required.

The previous meta-analysis was significantly limited by the small sample size and low power of the results. Additionally, the fixed effects model of statistical analysis was used despite significant heterogeneity in the included studies concerning the individual regimens, populations, and sample sizes. The fixed effects model tends to give greater weight to studies with larger sample sizes, thus undermining the effects of small sample size studies [24]. Also, it is not reasonable to assume that the intervention effects across all studies are identical, as the analysis demonstrates differences in the individual study effects [25]. We believe the sample sizes and interventional effects between the studies are adequately different; hence, a random effects model should be used. There was significant antibiotic resistance reported in one of the studies within the region being studied. [17,26]. This can hinder the comparability of results across studies, as other studies did not report similar resistance in their regions. Moreover, the study by Etik Ozer et al. reported two different pairs of 10-day and 14-day BQT, which were pooled together into a single intervention and control groups [16]. Our study assessed the two pairs separately.

Our study includes data from seven RCTs, providing a robust sample size and enhancing the reliability of the results. Moreover, by focusing on treatment-naive individuals, the study ensures that the results are not influenced by prior treatments and associated antibiotic resistance. Incorporating the latest RCTs, our study provides up-to-date evidence, reflecting the most recent clinical practices and findings in the treatment of *H. Pylori*. Moreover, by including studies from diverse geographic regions with larger sample sizes, our findings are more generalizable and applicable to a wider range of populations.

The non-inferiority of the 10-day BQT group demonstrated by our study also has logistical benefits. It reduces the wastage of drugs required for disease treatment while also reducing out-of-pocket costs for patients who would have to pay more for the longer duration of therapy [27]. This becomes even more important in the context of developing countries, which face medication shortages on a larger scale [28].

Clinicians may prescribe either the 10-day or 14-day BQT regimen, with knowledge of both treatments having comparable efficacy. This flexibility allows personalized treatment plans tailored to individual patient needs and circumstances. With no significant difference in outcomes, healthcare providers can optimize resource use by selecting the most appropriate regimen based on factors such as cost, availability, and patient preference without compromising treatment success. Our findings highlight the need for continued research on long-term outcomes and recurrence rates.

Another cost-effective method with a high eradication rate is sequential therapy, which has only recently been used in the context of quadruple therapy. It offers a high eradication rate with significant cost-effectiveness, which should be investigated further in large sample-size RCTs [29,30].

## Limitations

This study has some limitations, mainly the lack of longer follow-up times, which hinders the assessment of re-infection or long-term adverse events. Most of the studies had biases related to the randomization process (mainly the concealment of allocation) or selection bias. All the trials were single-center studies; such studies have limited external validity of the results and may overestimate the effect of the intervention. [31,32]

## Conclusion

Our meta-analysis comparing the 10-day and 14-day BQT regimens for treatment-naive *H. Pylori* infections found no significant difference in eradication rates, compliance, or incidence of adverse events between the two treatment durations. These findings suggest that both regimens are equally effective and safe for *H. Pylori* eradication, providing flexibility in treatment choices based on patient preferences, logistical constraints, and clinical circumstances.

## Abbreviations

BQT: Bismuth-containing quadruple therapy
ITT: Intention-to-treat
PP: Per protocol
UBT: Urea breath test

## Data Availability

All data produced in the present study are available upon reasonable request to the authors.

