## Supplementary File 1 for "Efficacy and safety of 10-day versus 14-day bismuth-containing quadruple therapy for H. pylori eradication: A systematic review and meta-analysis"

| **Database** | **Search Strategy** |
| --- | --- |
| **PubMed** | ("Helicobacter pylori"[Mesh] OR "h. pylori") AND ("Bismuth quadruple therapy" OR (("10-day bismuth quadruple therapy" OR"10-day bismuth-containing quadruple therapy" OR "10-day BQT") AND ("14-day bismuth quadruple therapy" OR"14-day bismuth-containing quadruple therapy" OR "14-day BQT"))) |
| **Cochrane** | ("helicobacter pylori" OR "h. pylori") AND ("bismuth-containing quadruple therapy" OR "bismuth quadruple therapy" OR "bismuth therapy") AND ("10-day") AND ("14-day") |
| **ClinicalTrials.gov** | “Helicobacter pylori” AND “Bismuth-containing therapy” |

**Table S1: Detailed Search Strategy and Results**

| **Study ID** | **Location** | **Included study arms** | **Included Participants** | **Mean Age** ± **SD in years** | **Male %** | **Duration of Follow-up** | **Confirmation of Diagnosis** | **Bismuth containing regimen** |
| --- | --- | --- | --- | --- | --- | --- | --- | --- |
| **Chen 2020** | China | 10-day vs 14-day Rabeprazole BQT | 120 vs 120 | 45.89 ± 12.92 vs 48.11 ± 12.39 | 56.7 vs 50.8 | 4 weeks | Rapid urease test, histology, culture, 14C‐urea breath test (14C‐UBT), or13C‐urea breath test (13C‐UBT) | 20 mg of rabeprazole, 1000 mg of amoxicillin, 500 mg of clarithromycin, and 220 mg of bismuth potassium citrate (BACPPI), twice daily for 10 days and 14 days |
| **Dore 2011** | Italy | 10-day vs 14-day Bismuth, Tetracycline, Metronidazole, and Esomeprazole | 215 vs 202 | 52 vs 53 | 37.7 vs 35.6 | 4-6 weeks | Endoscopy/Biopsy | 20 mg, tetracycline 500 mg, metronidazole 500 mg, and bismuth subcitrate caplets 240 mg, all twice daily |
| **Etik Ozer 2019** | Turkey | 10-day vs 14-day bismuth-containing quadruple treatment group (10-BQT vs 14-BQT groups)  10-day vs 14-day moxifloxacin-bismuth combined treatment groups (10-MBCT vs 14-MBCT groups) | 54 vs 54  54 vs 54 | 40 ± 11.0 vs 44 ± 12.5  41 ± 11.5 vs 43 ± 11.5 | 31.5 vs 35.2  40.7 vs 42.6 | 6-12 weeks | Gastric Biopsy | Regimen 1 (BQT): PPI (esomeprazole 40 mg bid), bismuth subsalicylate (524 mg bid), metronidazole (500 mg tid), and tetracycline (500 mg qid) for (10 vs 14) days Regimen 2 (MBCT): PPI (esomeprazole 40 bid), amoxicillin (1000 mg bid), and moxifloxacin (500 mg qd) and bismuth subsalicylate (524 mg bid) |
| **He 2023** | China | 10-day vs 14-day Antofloxacin containing BQT (ANT10 vs ANT14) | 395 vs 387 | 43.0 ± 11.8 vs 44.2 ± 11.1 | 51.6 vs 55.3 | 6 weeks | C-UBT positive, endoscopy/biopsy | Colloidal bismuth pectin 200 mg t.i.d., Esomeprazole 20 mg b.i.d., Amoxicillin 1 g b.i.d., and Antofloxacin 200 mg q.d. |
| **Lu 2023** | China | 10-day vs 14-day Vonoprazan BQT | 78 vs 78 | 37.1 ± 19.7 vs 35.9 ± 11.5 | 41.0 vs 47.4 | 6-8 weeks | Gastric biopsy using histochemical staining, tissue culture, the 14C-urea breath test (UBT), and/or the 13C-UBT. | vonoprazan 20 mg daily, amoxicillin 1000 mg, furazolidone 100 mg, and colloidal bismuth 200 mg twice a day |
| **Niu and Bai 2022** | China | 10-day vs 14-day Ilaprazole, Amoxicillin. Furazolidone, Bismuth Glycyrrhizinate (10-IAFB vs 14-IAFB) | 150 vs 150 | 44.25 ± 12.67 vs 44.10 ± 13.90 | 45.5 vs 42.1 | 4-6 weeks | C13 Urea Breath Test | 5 mg of Ilaprazole twice a day, 220 mg of bismuth glycyrrhizinate twice a day, 1.0 g of amoxicillin twice a day, and 0.1 g of furazolidone twice a day for 10 & 14 days |
| **Yang 2024** | Taiwan | 10-day vs 14-day Bismuth Quadruple Therapy (10-BQT vs 14-BQT) | 157 vs 156 | 55.0 ± 14.6 vs 55.0 ± 13.0 | 52.9 vs 42.9 | At least 4 weeks | 1) Histological evidence of H.pylori infection from gastric biopsy. 2) A Positive Rapid urease test. 3) A Positive C13 Urea Breath Test | Esopmeorazole 40 mg twice daily, Colloidal Bismuth Subcitrate 120 mg four daily, Metronidazole 500 mg thrice daily, Tetracycline 500 mg four time daily |

**Table S2: Detailed study characteristics (b.i.d. = Twice daily dose, t.i.d. = Three times a day)**
